# Troponin Level at Presentation as a Prognostic Factor Among Patients Presenting with non-ST Segment Elevation Myocardial Infarction

**DOI:** 10.1101/2023.04.27.23289237

**Authors:** the Jerusalem platelets thrombosis and intervention in cardiology (JUPITER-18) study group, Ranel Loutati, Nimrod Perel, Sharon Bruoha, Louay Taha, Meir Tabi, David Marmor, Itshak Amsalem, Rafael Hitter, Mohammed Manassra, Kamal Hamayel, Hani Karameh, Tommer Maller, Yoed Steinmetz, Mohammad Karmi, Mony Shuvy, Michael Glikson, Elad Asher

## Abstract

**Introduction:** Timely reperfusion within 120 minutes is strongly recommended in patients presenting with non-ST segment myocardial infarction (NSTEMI) with very high-risk features. Evidence regarding the use of high sensitivity cardiac troponin (hs-cTn) concentration upon admission for the risk-stratification of patients presenting with NSTEMI in order to expedite percutaneous coronary intervention (PCI) and thus potentially improve outcomes is limited.

**Methods:** All patients admitted to a tertiary care center intensive cardiac care unit (ICCU) between July 2019 – July 2022 were included and were followed for up to 3 years. Hs-cTnI level on presentation was recorded and patients were divided into four quartiles according to their hs-cTnI level on admission. Association between the initial hs-cTnI level and all-cause mortality during the follow-up period was examined.

**Results:** A total of 544 NSTEMI patients with a median age of 67 were included. There was no difference between the initial hs-cTnI level groups regarding age and comorbidities. A higher mortality rate was observed in the highest hs-cTnI quartile as compared with the lowest hs-cTnI quartile (16.2% vs. 7.35%, p=0.03) with Hazard ratio (HR) for mortality of 2.6 (CI: 1.23-5.4; p=0.012).

**Conclusions:** Patients presenting with NSTEMI and higher Hs-cTnI levels on admission were at higher risk for mortality during follow-up. This finding supports further prospective studies to examine the impact of early reperfusion strategy on mortality in patients presenting with NSTEMI according to degree of troponin elevation on admission.

## Introduction

Although substantial progress has been made in the invasive and pharmacologic therapies of acute coronary syndromes (ACS), the burden of ischemic heart disease is growing with more than 7 million people diagnosed annually with ACS worldwide ^1^. Timely invasive reperfusion is the mainstay of treatment and is associated with a mortality reduction of up to 9% in ST-segment elevation myocardial infarction (STEMI) and up to 6.5% in non ST-segment elevation myocardial infarction (NSTEMI) ^1,2^. However, while immediate revascularization is strongly recommended for patients presenting with STEMI, patients presenting with NSTEMI undergo risk stratification to determine the timing for coronary angiography and reperfusion ^3,4^. Urgent revascularization (< 2 hours) is recommended in the setting of NSTEMI with very high-risk features (i.e. hemodynamic instability, recurrent chest pain, malignant arrythmia, pulmonary congestion related to the MI, and cardiogenic shock). Otherwise, coronary angiography within 24 hours is advised. Cardiac biomarkers, in particular high-sensitivity cardiac troponin (hs-cTn), have shown promise in risk assessment of ACS patients. Hs-cTn has predictable kinetics in the setting of acute MI, and peak levels correlate with both ischemic time and quantity of jeopardized myocardium ^5^. Studies have consistently shown that elevated hs-cTn has high and incremental prognostic value in NSTEMI ^6-10^. In contrast, outcome data is inconsistent when hs-cTn levels are mildly elevated and/or non-dynamic ^9^. Current guidelines assign a class I level of recommendation for immediate and serial hs-cTn measurements for risk and prognostic stratification in NSTEMI ^4^, and urgent reperfusion is recommended in very high risk NSTEMI patients ^11,12^. However these guidelines do not rely on hs-cTn levels to determine whether the urgent invasive approach is indicated. Hence, the aim of the current study was to evaluate whether high level of hs-cTn I (hs-cTnI) on admission correlates with outcomes.

## Methods

### Study population

All patients diagnosed with NSTEMI who were admitted to a tertiary care intensive cardiac care unit (ICCU) at Shaare Zedek Medical Center between July 2019 – July 2022 were prospectively recruited.

The diagnosis of NSTEMI was based on symptoms of myocardial ischemia, new ECG ischemic changes, and a rising and/or falling pattern of high-sensitivity troponin with at least one value above the 99^th^ percentile URL, according to the ESC guidelines for ACS ^4^

### Data collection

Data were anonymously documented in the ICCU by the local coordinator and prospectively submitted into an electronic case report form (eCRF). Data were checked for accuracy and out-of-range values by the coordinating unit. Demographic data, presenting symptoms, comorbid conditions, and physical examination were systematically recorded. Laboratory, imaging, angiographic results, and clinical course data were collected as well.

Patients were followed for up to three years after presentation. Patients were divided into four quartiles according to their hs-cTnI level on admission. The first quartile served as the reference group. Hs-cTnI assay was determined in a central laboratory (ARCHITECT STAT hs-cTnI immunoassay) with a 99^th^ percentile reference level of 17 ng/L for female, 35 ng/L for male and 28 ng/L for overall ^13^. Invasive and pharmacologic treatments were in accordance with the European society of cardiology (ESC) guidelines for ACS^13^.

The Institutional review board approved the study based on strict maintenance of participants anonymity by de-identifying during database analysis. No individual consent was obtained. Moreover, the authors have no conflicts of interest to declare. No funding was applied to the study. All methods were performed in accordance with the relevant guidelines and regulations.

### Study Outcomes

The primary outcome was all-cause mortality with a follow-up time of up to 3 years. Overall mortality rate was determined from the Israeli Ministry of Internal Affairs.

### Statistical analysis

Continuous variables were expressed as mean ± standard deviation if normally distributed or median with interquartile range if skewed. Categorical variables were presented as frequency (%). Continuous data were compared with the Student’s t-test and Mann-Whitney test for comparison of normally and non-normally distributed continuous variables, respectively. Categorical data were compared with the use of the chi-square test or Fisher exact test. For survival analysis patients were censored in the case of death or diagnosis of malignant cancer during follow-up.

The probability of death according to the study groups was graphically displayed according to the method of Kaplan–Meier, with a comparison of cumulative survival across strata by the log-rank test. Univariate Cox proportional hazards regression modeling was used to determine the unadjusted Hazard Ratio (HR) for all-cause mortality of patients with high presenting troponin levels. All analyses were performed R software version 3.4.4 (R Foundation for Statistical Computing). An association was considered statistically significant for a two-sided P value of less than 0.05.

## Results

### Baseline characteristics

A total of 665 patients with NSTEMI were screened, of them 121 were excluded due to missing data or loss of follow-up, hence, the final study population included 544 patients. Mean age was 68 (±11) years old and 122 (22%) were female. Patients’ characteristics are present in table 1. Three hundred and fifty (64%) patients suffered from hypertension, 233 (43%) from diabetes mellitus (DM), 342 (63%) from dyslipidemia, 184 (34%) patients were active smokers, and 39 (7%) patients had prior ischemic heart disease.

**Table 1.**
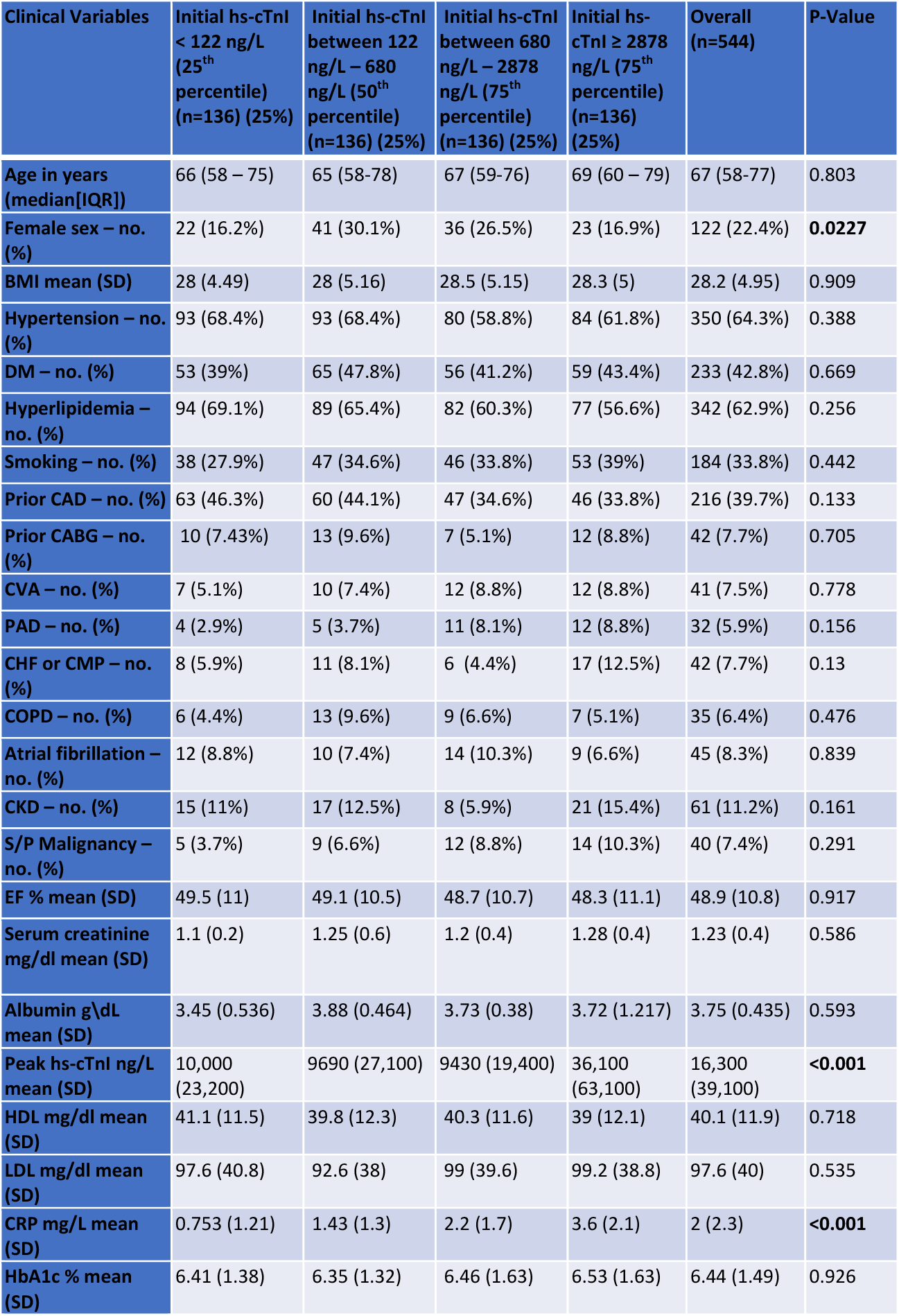
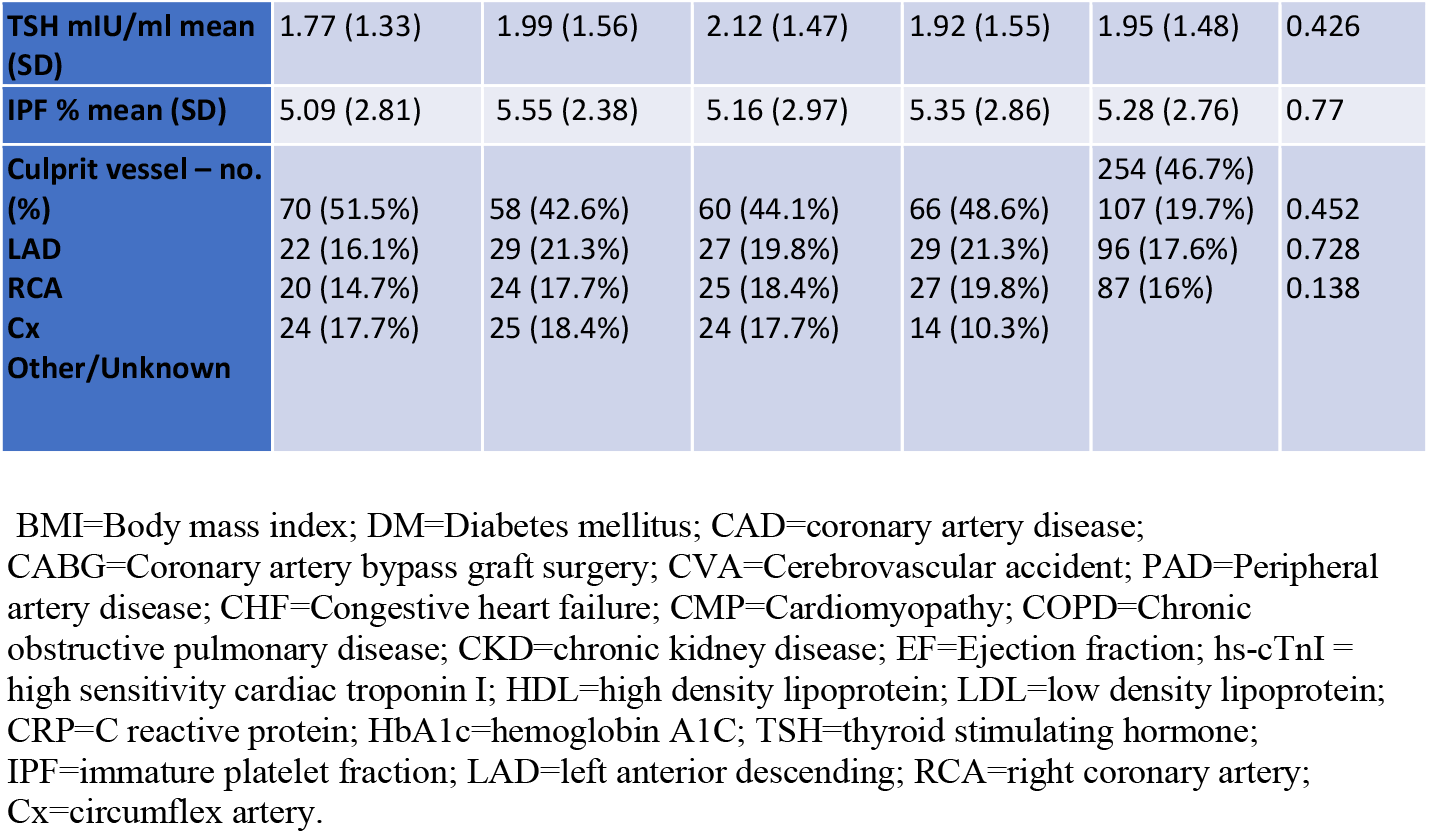
Baseline characteristics

### Hs-cTnI levels at presentation

Hs-cTnI levels in each quartile were: a) ≤122 ng/L; b) 123-680 ng/L; c) 681-2877 ng/L and d) ≥ 2878 ng/L. Median initial hs-cTnI level was 680 ng/L (range 0-150,008 ng/L). There were no differences regarding age or comorbidities between the 4 initial hs-cTnI level quartiles. Prevalence of male gender, higher peak level of hs-cTnI during admission, C reactive protein (CRP), and fibrinogen were greater in the 4^th^ quartile as compared with the 1^st^ quartile (table 1).

### Interventions and Complications during Admission

Specific procedures performed during the ICCU admission course are reported in Table 2. PCI was performed in 369 (68%) patients, coronary angiography without intervention was performed in 119 (22%), coronary artery bypass grafting (CABG) was performed in 27 (4.9%) patients, and medication therapy alone was assigned to only 29 (5.1%) of the patients. There were no differences in the treatments between the patients in all quartiles as shown in Table 2. Overall complication rate during admission was 15% with no significant differences between the quartiles, as presented in Table 3. Acute renal failure (ARF) and shock were more common in the 4^th^ quartile as compared with the 1^st^ quartile (7.4% vs. 2.9%, p=0.07 and 9.5% vs. 5.1%, p=0.062, respectively).

**Table 2.** Interventions during admission

| **Type of procedure** | **Initial hs-cTnI < 122 ng/L (25<sup>th</sup> percentile) (n=136) (25%)** | **Initial hs-cTnI between 122 ng/L – 680 ng/L (50<sup>th</sup> percentile) (n=136) (25%)** | **Initial hs-cTnI between 680 ng/L – 2878 ng/L (75<sup>th</sup> percentile) (n=136) (25%)** | **Initial hs-cTnI ≥ 2878 ng/L (75<sup>th</sup> percentile) (n=136) (25%)** | **Overall (n=544)** | **P-Value** |
| --- | --- | --- | --- | --- | --- | --- |
| **PPCI – no. (%)** | 17 (12.5%) | 9 (6.6%) | 16 (11.8%) | 8 (5.9%) | 50 (9.2%) | 0.221 |
| **PCI – no. (%)** | 72 (52.9%) | 86 (63.2%) | 80 (58.8%) | 81 (59.6%) | 319 (58.6%) | 0.549 |
| **Cath. – no. (%)** | 31 (22.8%) | 29 (21.3%) | 31 (22.8%) | 28 (20.6%) | 119 (21.9%) | 0.99 |
| **CABG – no. (%)** | 8 (5.9%) | 4 (3%) | 8 (5.9%) | 7 (5.1%) | 27 (5%) | 0.681 |
| **Medications alone – no. (%)** | 8 (5.9%) | 8 (5.9%) | 1 (0.7%) | 12 (8.8%) | 29 (5.1%) | 0.669 |
| **Pacemaker/ICD – no. (%)** | 3 (2.2%) | 0 (0%) | 2 (1.5%) | 1 (0.7%) | 6 (1.1%) | 0.498 |
| **CPR – no. (%)** | 4 (2.9%) | 1 (0.7%) | 2 (1.5%) | 2 (1.5%) | 9 (1.7%) | 0.709 |
| **Mechanical Ventilation – no. (%)** | 8 (5.9%) | 5 (3.7%) | 4 (2.9%) | 5 (3.7%) | 22 (4%) | 0.79 |
| **RRT – no. (%)** | 4 (3%) | 9 (6.6%) | 7 (5.1%) | 7 (5.1%) | 27 (5%) | 0.432 |
| **IABP – no. (%)** | 2 (1.5%) | 2 (1.5%) | 2 (1.5%) | 5 (3.7%) | 11 (2%) | 0.654 |

PPCI=Primary Percutaneous Coronary Intervention; PCI=Percutaneous Coronary Intervention; Cath.=Coronary angiography without intervention; CABG=Coronary Artery Bypass Graft; ICD=Implantable Cardioverter Defibrillator; CPR=Cardio-Pulmonary Resuscitation; RRT=Renal replacement therapy; IABP=Intra-Aortic balloon pump

**Table 3.** Complications during admission

| **Type of complication** | **Initial hs-cTnI < 122 ng/L (25<sup>th</sup> percentile) (n=136) (25%)** | **Initial hs-cTnI between 122 ng/L – 680 ng/L (50<sup>th</sup> percentile) (n=136) (25%)** | **Initial hs-cTnI between 680 ng/L – 2878 ng/L (75<sup>th</sup> percentile) (n=136) (25%)** | **Initial hs-cTnI ≥ 2878 ng/L (75<sup>th</sup> percentile) (n=136) (25%)** | **Overall (n=544)** | **P-Value** |
| --- | --- | --- | --- | --- | --- | --- |
| **Malignant Arrhythmia – no. (%)** | 2 (1.5%) | 0 (0%) | 1 (0.7%) | 2 (1.5%) | 5 (0.9%) | 0.695 |
| **Shock\* – no. (%)** | 7 (5.1%) | 6 (4.4%) | 6 (4.4%) | 13 (9.5%) | 32 (5.9%) | 0.062 |
| **Stroke/TIA – no. (%)** | 0 (0%) | 1 (0.7%) | 3 (2.2%) | 1 (0.7%) | 5 (0.9%) | 0.695 |
| **ARF – no. (%)** | 4 (2.9%) | 2 (1.5%) | 3 (2.2%) | 10 (7.4%) | 19 (3.5%) | 0.07 |
| **Significant bleed – no. (%)** | 0 (0%) | 1 (0.7%) | 3 (2.2%) | 5 (3.7%) | 9 (1.7%) | 0.155 |

ARF=Acute Renal Failure
\*any type of shock

### Mortality rate during follow-up

Death rate was 10 (7.3%), 24 (17.6%), 14 (10.3%) and 22 (16.2%) for each quartile, respectively as shown in Figure 1. Age, left ventricular ejection fraction (LVEF), ischemic heart disease, heart failure, hypertension, and DM were all predictors of poor survival (Table 4).

**Table 4.** Univariate cox model

| Clinical variables | Hazard Ratio (HR) | 95% CI | P value |
| --- | --- | --- | --- |
| Age | 1.05 | 1.02-1.07 | <0.001 |
| Sex (Male) | 0.65 | 0.39-1.08 | 0.1 |
| BMI (continuous) | 0.99 | 0.94-1.04 | 0.7 |
| Obesity (BMI>30) | 0.9 | 0.54-1.50 | 0.7 |
| LVEF | 0.94 | 0.91-0.97 | <0.001 |
| Low-LVEF (<41%) | 2.87 | 1.75-4.70 | <0.001 |
| **Initial Troponin >= Q1 (122)** | 2.05 | 1.05-4 | 0.003 |
| **PCI** | 0.48 | 0.29-0.81 | <0.001 |
| Ischemic heart disease | 2.90 | 1.56-5.40 | <0.001 |
| Heart failure | 2.78 | 1.49-5.20 | 0.001 |
| Arterial hypertension | 2.71 | 1.42-5.17 | 0.002 |
| Diabetes mellitus | 1.68 | 1.04-2.70 | 0.032 |
| COPD | 1.40 | 0.66-2.94 | 0.4 |
| Dyslipidemia | 1.30 | 0.79-2.14 | 0.3 |
| Cerebrovascular accident | 1.33 | 0.61-2.91 | 0.5 |
| Peripheral vascular disease | 2.41 | 1.23-4.71 | 0.01 |
| Smoking | 0.46 | 0.26-0.83 | 0.01 |
| Malignancy | 1.23 | 0.56-2.68 | 0.6 |

BMI=Body Mass Index; LVEF=Left ventricle ejection fraction; PCI=Percutaneous Coronary Intervention; COPD = Chronic Obstructive Pulmonary Disease

**Figure 1.**
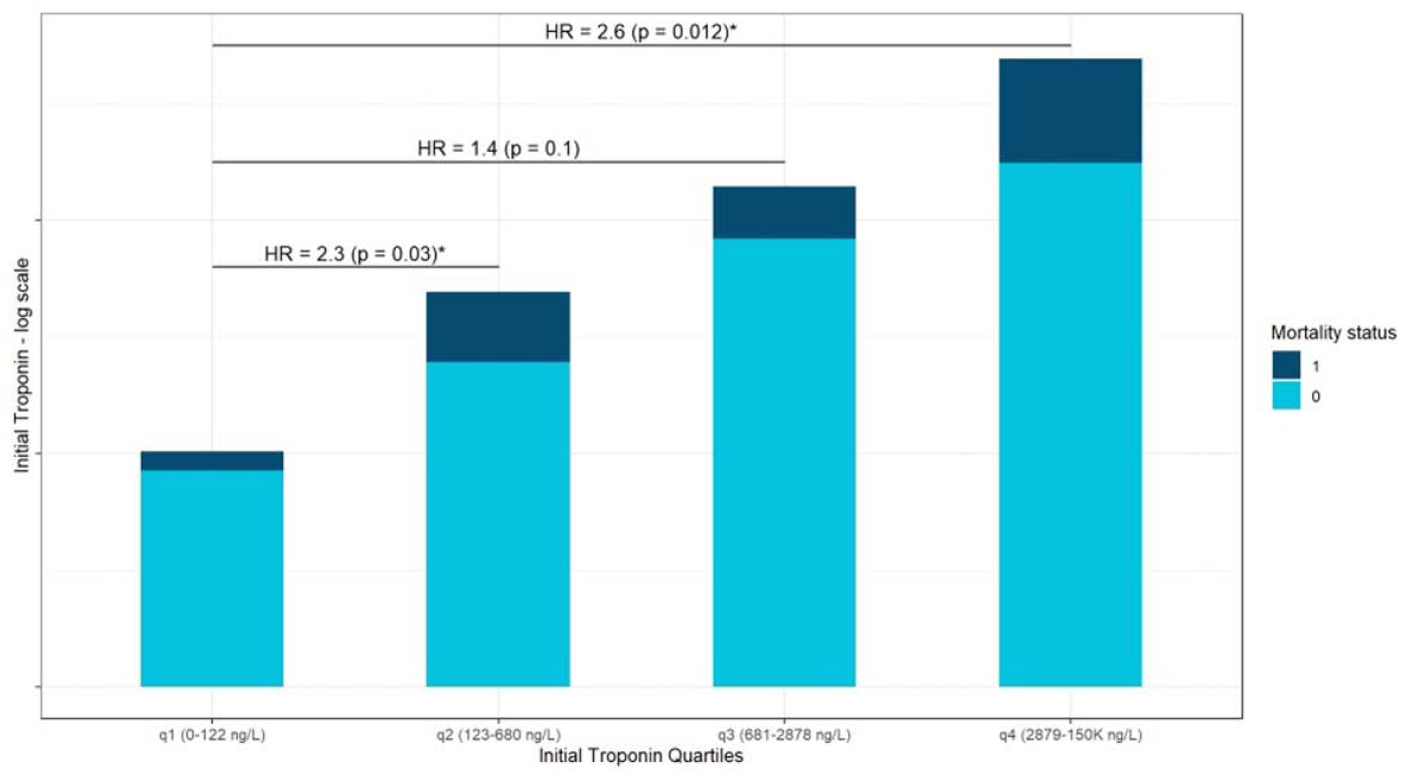
Bar plot for the four groups of patients with initial hs-cTnI at various quartiles: This bar plot shows the relative portion of deceased patients in each of the four initial hs-cTnI groups of patients. Hazard ratios (HR) for mortality between the 1^st^ quartile and the 3 others are also depicted, indicating greater risk of mortality as initial hs-cTnI rises.

The cumulative probability of death at 1 year was 6.5±2.4%, 5±3.8%, 10±2.5%, and 22±4% for each quartile respectively (Figure 2; Log Rank p = 0.026 for overall difference during follow-up). Patients who presented with initial hs-cTnI in the 4^th^ quartile were 2.6 times more likely to die at 1 year follow-up, as compared with patients in the 1^st^ quartile (HR 2.6 95% CI 1.23 – 5.4, p=0.012). A multivariate Cox model adjusted for age, gender, and serum creatinine showed consistent results (HR 2.26 95% CI 1.04 – 4.89, p=0.04) as was further adjustment of the model for revascularization (PCI and CABG) during admission (HR 2.15 95% CI 0.95-4.65, p=0.06).

**Figure 2.**
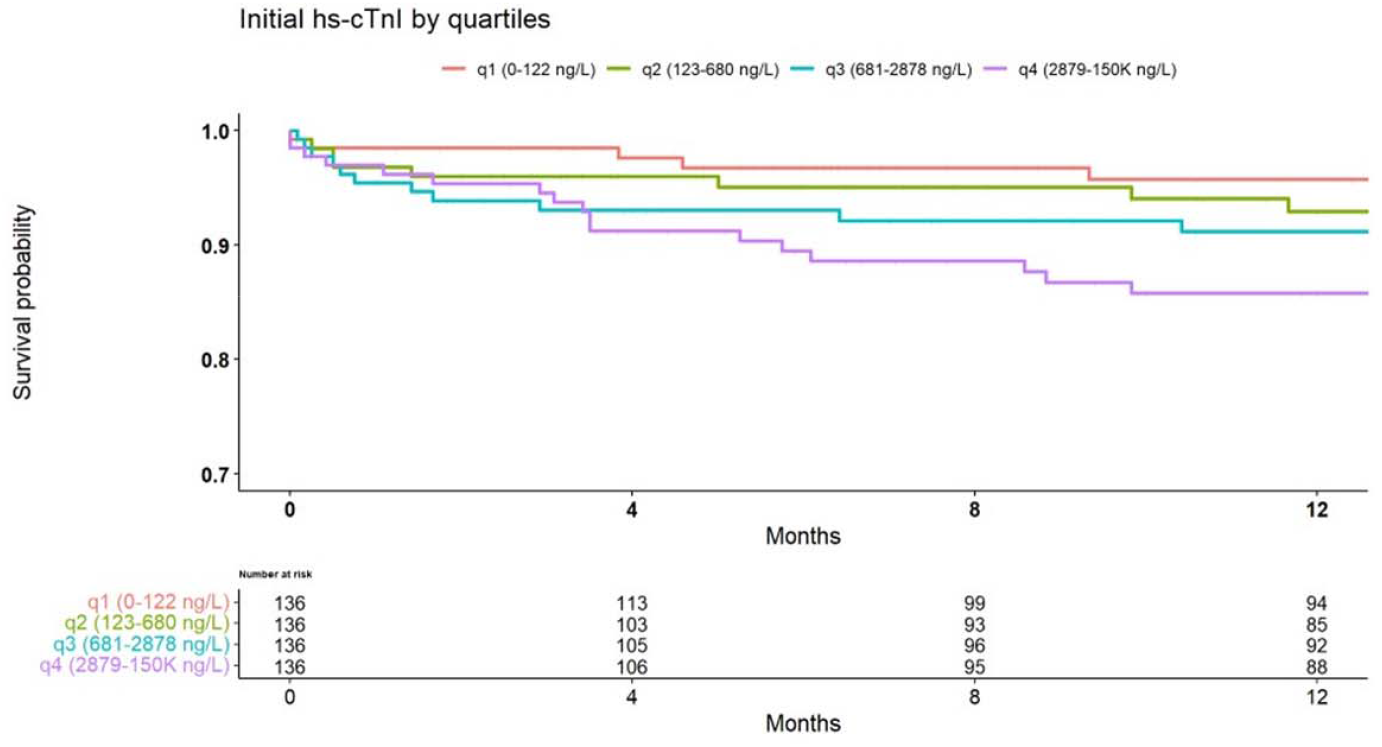
Kaplan-Meier curves for the four mutually exclusive groups based on initial hs-cTnI quartiles: Kaplan-Meier survival curves demonstrating lower survival probability in patients with higher initial hs-cTnI levels. Log Rank p < 0.001

### Dose response effect of initial hs-cTnI levels on mortality

To investigate the potential dose response relationship between initial hs-cTnI and mortality, we plotted the risk of death at 1 year of follow-up using Kaplan-Meier survival analysis across various levels of initial hs-cTnI, as depicted in Figure 3. The plot exhibits a sigmoidal curve, characterized by a gradual increase in mortality with rising levels of initial hs-cTnI, until reaching a plateau at hs-cTnI = 1000 ng/L. Notably, a second peak in mortality risk emerges at hs-cTnI = 2878 ng/L.

**Figure 3.**
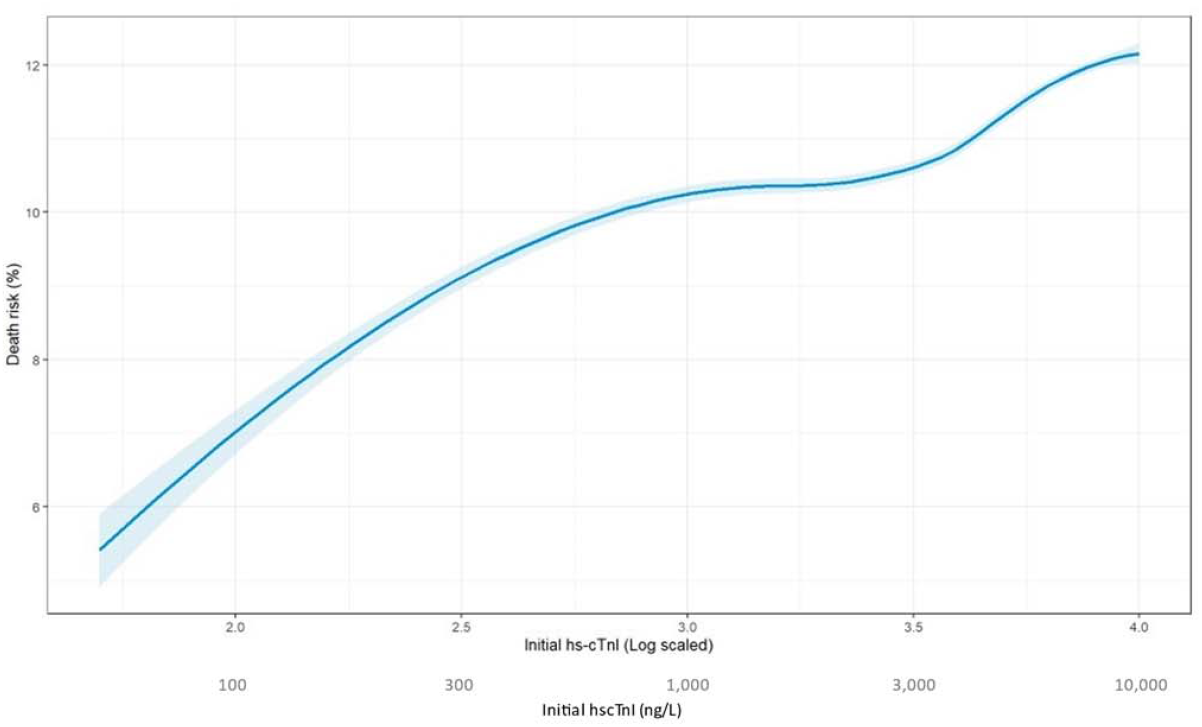
Interpolated Death Risk (%) based on survival probability at various hs-cTnI values, demonstrating later peak in mortality at hs-cTnI = 2878 ng/L:This graph represented the interpolated risk for mortality at 1 year of presentation, based on initial hs-cTnI levels. As initial hs-cTnI levels increased, so is the risk for mortality. A second, later peak, in the death risk is seen when initial hs-cTnI levels reached the 4^th^ quartile.

## Discussion

The main findings of our study are a) we have demonstrated that any increase in hs-cTnI levels upon admission for patients with NSTEMI carries prognostic implications, b) we have identified a subgroup of patients with initial high hs-cTnI levels representing the 4th quartile of our study (hs-cTnI ≥ 2878 ng/L), who are at higher risk of mortality, and c) even after adjusting the multivariate analysis, initial high hs-cTnI levels remained an independent predictor for 1-year mortality.

To the best of our knowledge, this study is the first to examine the association between hs-cTnI levels upon hospital admission and long-term overall survival in patients diagnosed with NSTEMI. Prior studies that have examined the association between initial troponin levels and mortality have included patients with a broader range of diagnoses, encompassing all types of acute myocardial infraction ^14^ or patients presenting with chest pain^15^. Another significant distinction between our study and previous ones is our focus on examining troponin levels at presentation, as opposed to peak troponin levels or changes in troponin during admission course, which has been linked to adverse clinical outcomes and mortality^15-17^.

Our first finding further supports the conclusions of Antman et al. regarding the role of troponin level upon hospital admission and short-term mortality in ACS patients^18^. The identification of a higher risk subgroup with hs-cTnI levels greater or equal to 2878 ng/L is very intriguing, as this subgroup did not exhibit any distinctive baseline characteristics or complications. We hypothesized that the heightened risk of mortality in those patients is likely a result of later changes that occur in the myocardium of NSTEMI patients, such as remodeling, which is related to elevated risk of death^19^. Hence there were no between-group differences in the short-term period. In a large observational study comprising 4,123 patients with non-ST-segment elevation ACS, Kontos et al. demonstrated a gradual rise in cardiac mortality as peak cTnI levels increased at 30-day and 6-month follow-up intervals^15^. Our results are consistent with these findings. However, our aim was to investigate specifically the association between initial cTnI levels and mortality over a longer follow-up period. From a clinical perspective, our results might have greater clinical implications due to the dynamic nature of ACS patients.

Bagai et al. showed that there is a differential relationship between the magnitude of peak troponin elevation and long-term mortality in ACS patients treated with and without revascularization^16^. Our findings also suggest better prognosis after revascularization regarding the association between initial hs-cTnI levels and mortality, although this was not the aim of our study and it was not powered for that. he association between initial hs-cTnI levels and mortality, and the lower mortality rate in revascularized patients, imply that initial hs-cTnI levels may be useful in determining the need for urgent revascularization in NSTEMI patients. Current NSTEMI guidelines advise revascularization within 120 minutes in patients with very high-risk characteristics^4^, regardless of the level of cardiac biomarkers. However, based on our results and previous critical studies, initial hs-cTnI can significantly add to the decision-making process for urgent revascularization among NSTEMI patients.

## Study limitations

Our study has several limitations: 1) The study was conducted in a single tertiary-care ICCU, resulting in a relatively small study population of 544 patients. Nevertheless, we included all-comers in a real-life setting. 2) Our analysis was based on overall mortality rather than cardiovascular mortality. Though mortality statistics in Israel closely resemble those of the European Union, where cardiovascular death is the second most prevalent cause of death following cancer^20^. 3) There may be unmeasured clinical variables that could explain our observations, including the time elapsed between symptom onset to hs-cTnI testing, details regarding coronary angiography beyond the culprit vessel, including lesion description and the exact type of stents employed in the PCI group.

## In conclusion

Despite the prognostic implications of troponin levels in ACS, current guidelines for urgent revascularization in NSTEMI patients do not incorporate this biomarker in their decision-making process. In our study, we have demonstrated an association between hs-cTnI levels upon admission and overall survival in patients diagnosed with NSTEMI. Furthermore, we identified a subgroup that is at a heightened risk of mortality according to their initial hs-cTnI level. Importantly, this association becomes less significant when early reperfusion strategy is taken into account. These findings suggest that initial hs-cTnI levels could serve as an important decision-making tool for determining reperfusion strategies in NSTEMI patients. Further prospective studies are warranted to investigate the impact of early reperfusion strategies on mortality in NSTEMI patients categorized by their initial presenting troponin levels.

## Data Availability

The datasets generated and/or analyzed during the current study are available from the corresponding author on reasonable request.

## Notes

### Competing Interest Statement

The authors have declared no competing interest.

### Funding Statement

No external funding was received

